# Automated interpretation of the coronary angioscopy with deep convolutional neural networks

**DOI:** 10.1101/19001693

**Authors:** Toru Miyoshi, Akinori Higaki, Hideo Kawakami, Osamu Yamaguchi

**Affiliations:** Department of Cardiology, Ehime Prefectural Imabari Hospital, Imabari, Japan; Department of Cardiology, Pulmonology, Hypertension and Nephrology, Ehime University Graduate School of Medicine, Toon, Japan; Hypertension and Vascular Research Unit, Lady Davis Institute for Medical Research, Sir Mortimer B. Davis Jewish General Hospital, McGill University, Montreal, Canada; Department of Cardiovascular Medicine, Osaka University Graduate School of Medicine, Suita, Japan

**Keywords:** coronary angioscopy, deep convolutional neural network, generative adversarial networks, artificial intelligence

## Abstract

**Background:** Coronary angioscopy (CAS) is a useful modality to assess atherosclerotic changes, but interpretation of the images requires expert knowledge. Deep convolutional neural networks (DCNN) can be used for diagnostic prediction and image synthesis.

**Methods:** 107 images from 47 patients, who underwent coronary angioscopy in our hospital between 2014 and 2017, and 864 images, selected from 142 MEDLINE-indexed articles published between 2000 and 2019, were analyzed. First, we developed a prediction model for the angioscopic findings. Next, we made a generative adversarial networks (GAN) model to simulate the CAS images. Finally, we tried to control the output images according to the angioscopic findings with conditional GAN architecture.

**Results:** For both yellow color (YC) grade and neointimal coverage (NC) grade, we could observe strong correlations between the true grades and the predicted values (YC grade, average r value = 0.80 ± 0.02, p-value <0.001; NC grade, average r value = 0.73 ± 0.02, p < 0.001). The binary classification model for the red thrombus yielded 0.71 ± 0.03 F1-score and the area under the ROC curve (AUC) was 0.91 ± 0.02. The standard GAN model could generate realistic CAS images (average Inception score = 3.57 ± 0.06). GAN-based data augmentation improved the performance of the prediction models. In the conditional GAN model, there were significant correlations between given values and the expert’s diagnosis in YC grade and NC grade.

**Conclusion:** DCNN is useful in both predictive and generative modeling that can help develop the diagnostic support system for CAS.

## Introduction

Coronary angioscopy (CAS) is a unique imaging device which enables direct visualization of the vessel lumen and provides comprehensive information about atherosclerotic changes (1, 2). It has been reported that CAS has a higher ability to detect neointimal vulnerability after stent implantation than other modalities (3). However, this procedure requires expert knowledge for interpretation and is therefore not common to general cardiologists. Currently, there is an increasing number of studies about the application of artificial intelligence in the field of cardiology (4, 5). In particular, deep convolutional neural networks (DCNN) have become popular in medical image analysis (6, 7) and its application is extended to the generative tasks such as generative adversarial networks (GAN) (8). Considering its ability of feature abstraction, we think DCNN can be used for the automated interpretation of CAS images and enhance the outreach of this procedure. In this study, we aimed to 1. develop a data-driven prediction model for CAS findings as a diagnostic support system and 2. synthesize realistic CAS images for educational purposes using GAN, that may in turn improve the performance of diagnostic system. Additionally, we demonstrated the outcome of conditional image synthesis according to the CAS findings.

## Methods

This retrospective observatory study was performed following the principles of the Declaration of Helsinki and the Japanese ethical guidelines for clinical research. All patients provided written informed consent. The study protocol was approved by the institutional review boards and the ethics committees of Ehime Prefectural Imabari Hospital. Part of the datasets, including script files will be available from the online repository (http://dx.doi.org/10.17632/9dx23j5d64.1).

### Image acquisition from patients

We retrospectively assessed 107 consecutive lesions after stent implantation using CAS for 47 patients who had undergone PCI between February 2014 and October 2017 at Ehime Prefectural Imabari Hospital. Detailed patient characteristics are indicated in Supplementary Table S1. Catheterization was performed with a radial, brachial or femoral approach using ≥6-F guiding catheters. CAS was performed with the FT-203F (FiberTech Co. Ltd., Tokyo, Japan) non-obstructive coronary angioscope system and the VISIBLE (FiberTech Co. Ltd.) optical fiber as previously reported (9).

### Evaluation of the angioscopy

Each CAS analysis was performed as agreed by 2 independent cardiologists. Neointimal coverage (NC) grade was evaluated using a 4-point grading scale, from 0 (no coverage) to 3 (complete coverage). Plaque yellow color (YC) was assessed using a 4-grade system, from 0 (white) to 3 (bright yellow). The presence of red thrombus was also assessed as previously described (10). Representative CAS images for each grade are shown in Fig 1.

**Fig 1.**
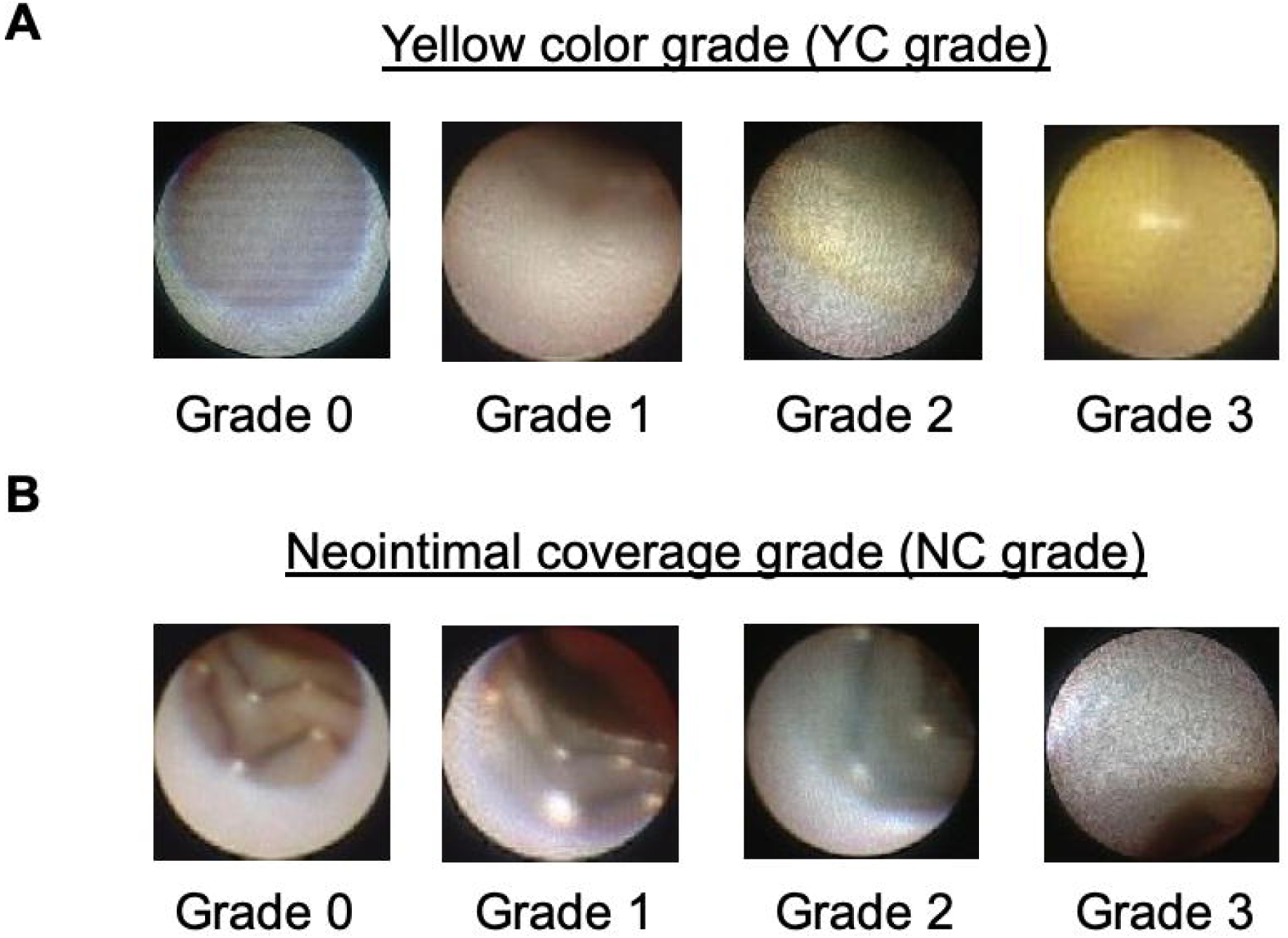
Definitions of CAS findings. Panel A shows the representative images for each yellow color (YC) grades. Panel B is for the neointimal coverage (NC) grades. All shown images are converted to 96×96 pixels.

### Additional image collection from MEDLINE-indexed articles

In order to address the class imbalance problem, we additionally collected angioscopic images using PubMed search. We selected 142 MEDLINE-indexed articles which included coronary angioscopy pictures, published between April 2000 and April 2019 (Supplementary Table S2). Pictures with excessive modifications or annotations (e.g. lines and arrows) were excluded from the analysis. All collected images were curated by at least two expert cardiologists. Since not all pictures were obtained from the stent implanted site, the NC grades were uniformly defined as 3 when stent struts were not observed.

### Image data preprocessing

All collected images were converted to the Joint Photographic Experts Group (JPEG) format with 24-bit color data and resized to 96×96 pixel size with the image processing tool of Python interpreter (Pillow; Alex Clark and Contributors). In reference to the previous report (11), we employed geometric data augmentation; rotation with 4 different angles. For the appropriate cross-validation, training and test datasets were properly separated, so as not to share the same image source.

### Structure of the prediction models

We designed a simple 4-layer DCNN model which can be universally used for the assessment of YC grade, NC grade and red thrombus. Mean squared error as used as a loss function for the regression tasks (predicting the grade scores). For the binary classification, we set sigmoid function as the final activation function and used binary cross entropy as a loss function. Adaptive moment estimation as used in the all prediction models as an optimization algorithm.

### Structure of the GAN models

We designed a standard deep convolutional GAN and a conditional GAN model, according to the original articles (12, 13). Fig 2 shows the schematic representation of the conditional GAN model in this study. All CAS findings were used as the conditioning information to train the model. For the standardization, YC and NC grades were divided by three to be ranged between 0 and 1. The conditioning vector was concatenated with a gaussian noise and passed to the generator network. For the discriminator network, conditions were combined with the image data as input and also passed to the fully connected layer.

**Fig 2.**
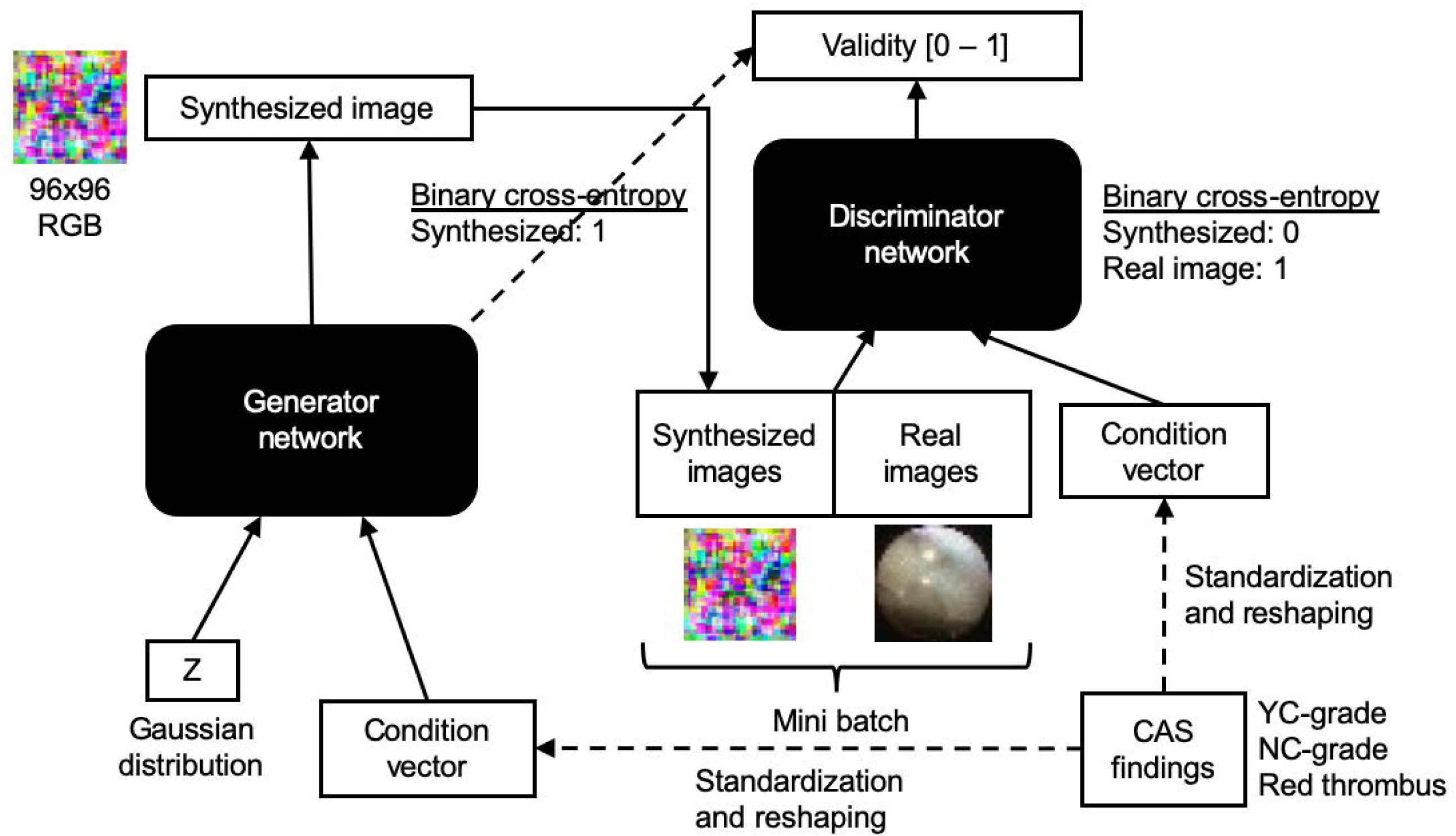
Schematic representation of the conditional GAN model. CAS findings are converted to condition vector and passed to both generator and discriminator networks.

### Evaluation method for the performance of GAN models

For the standard GAN model, Inception scores were calculated for a randomly selected 1000 images from the original data and for the 1000 generated images respectively. We employed a blinded visual scoring system for the evaluation of the conditional GAN model. Two experts were asked to grade the generated pictures without any information about the given conditions. Per each grade, 12 images were generated by the conditional GAN. These blinded scores were compared with the given conditions and the correlation coefficient was calculated.

### Method for the GAN-based data augmentation

In order to test the capability of GAN-based image synthesis as a data augmentation method, we split the dataset differently from the previous experiment. Namely, images from our hospital were used as a fixed validation dataset (n = 107) and the prediction model was trained on the online-collected images (n = 864) or on the augmented dataset where the original images were combined with the same number of synthesized images (n = 1728). To see the pure effect of this augmentation method, no geometrical transformation was conducted for this experiment. The standard GAN was trained only on the online-collected images to prevent the possible overfitting phenomenon. Model training was repeated 5 times with different random seeds and the mean performance indices were calculated.

### Statistical analysis

Data is presented as mean ± standard error. Five-fold cross validation was used to evaluate prediction models. For the assessment of regression models, we employed Pearson’s correlation coefficient (r-value) and mean absolute error (MAE) as indicators of the model performance. R-values higher than 0.7 were considered to show strong positive linear relationships.

For the binary classification task, we used precision, recall, specificity, negative predictive value and F1-scores as indicators. Receiver operator characteristics (ROC) curve and the area under the curve (AUC) were also analyzed. P-values less than 0.05 were regarded as statistically significant. All analysis was performed using SciPy module in the python library.

## Results

### Characteristics of the articles in the PubMed search

A total 864 different angioscopic images were obtained from 142 articles (Supplementary Table S2). Among these 142 articles, 82 (57.7%) were original articles and 44 (30.9%) were case reports. 12 review articles, 2 letters, a rapid communication and an editorial article were also included. Among them, 89 (62.7%) articles provide images of stented lesions, and the rest were about de novo lesions. On average, 7 pictures were available from an original article and 4.4 pictures from a case report. *Circulation Journal* (29 articles, 20.4%), *JACC Cardiovascular Intervention* (16 articles, 11.3%) and *International Journal of Cardiology* (9 articles, 6.3%) were the top 3 most frequently found journals through the online search.

### Distributions of CAS findings

In our hospital data, the most frequently observed finding was YC grade 0, NC grade 1 without any thrombus (17.8% of the total 107 images). YC grade 3 was not observed except in NC grade 3 without thrombus. Red thrombi were most frequently observed in YC grade 1, NC grade 1 area (Fig 2A). On the other hand, YC grade 0, NC grade 1 is the most frequent finding among the images collected through PubMed search (16.2% of the total 864 images). (Fig 2B). On the whole, lower NC grades were more frequently found in the published articles’ pictures. Distribution in the mixed data is shown in Fig 2C.

### Diagnostic prediction model for the angioscopic findings with DCNN

In both the YC grade and NC grade prediction model, we observed strong and significant correlations between the true grades and the predicted values (YC grade, average r value = 0.80 ± 0.02, p-value <0.001, average MAE = 0.16 ± 0.01; NC grade, average r value = 0.73 ± 0.02, p < 0.001, average MAE = 0.17 ± 0.01). Representative box-whisker plots are shown in Fig 3. The binary classification model for the red thrombus achieved 0.87 ± 0.04 in precision, 0.62 ± 0.06 in recall, resulting in 0.71 ± 0.03 F1-score. The specificity was 0.96 ± 0.01 and the negative predictive value was 0.89 ± 0.01. Average AUC was 0.91 ± 0.02, calculated from ROC curves of 5-fold cross validation (Supplementary Figure S1).

**Fig 3.**
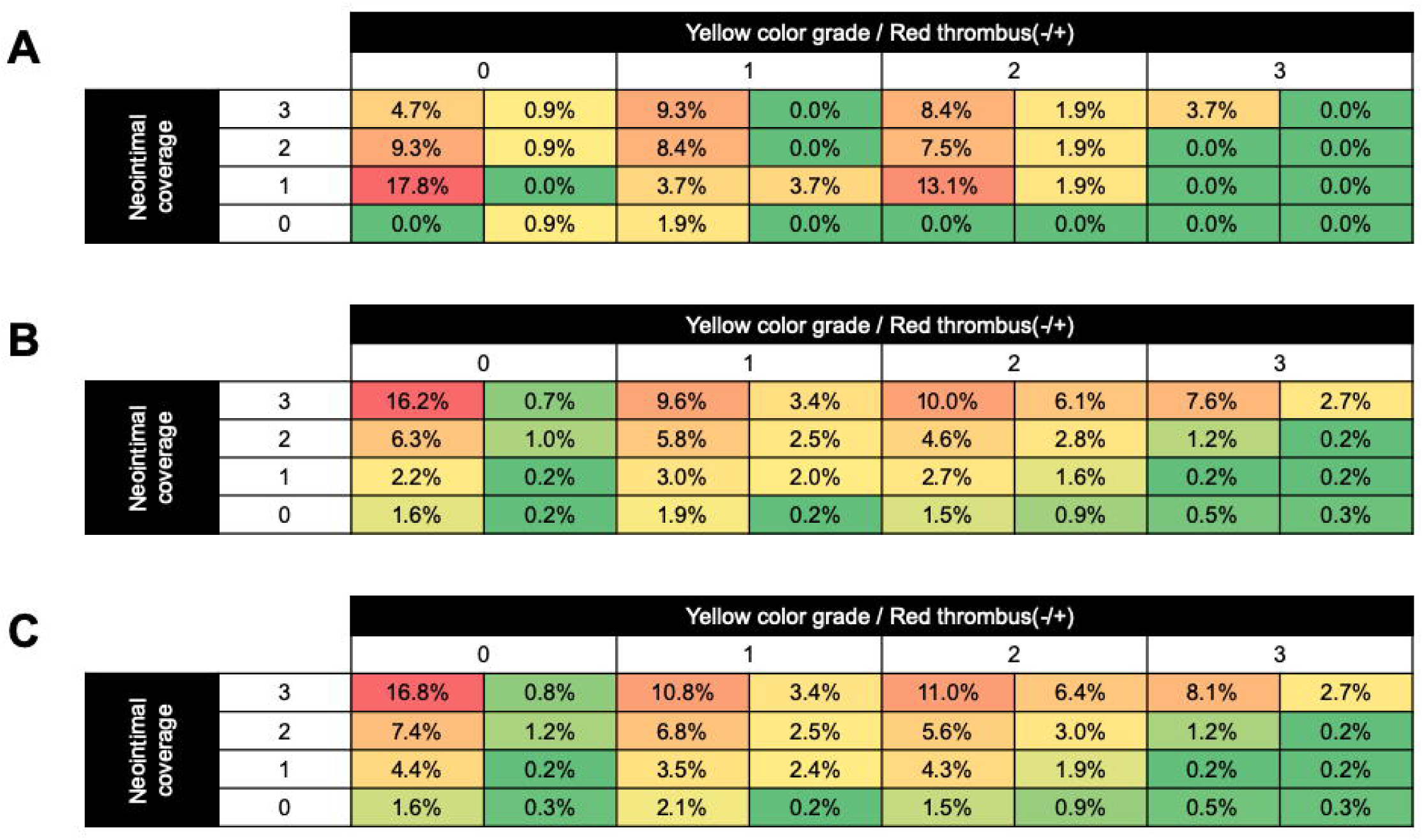
Heatmaps for the distribution of the angioscopic findings. The percentage of the population with the specific finding was shown for the images obtained from our hospital (A), PubMed search (B) and in the merged dataset (C). In each area of the matrix, the right sided cell indicates the population with red thrombus. Cells with higher percentage were highlighted with red background color as a heatmap (colors are assigned independently for each panel).

### Synthesized CAS images by the GAN models

The standard GAN model generated a variety of angioscopic images (Fig 4A) with the average Inception score of 3.57 ± 0.06. Images were visually realistic allowing us to do the annotation, but the score was significantly lower than the original dataset (3.90 ± 0.01, p<0.01).

**Fig 4.**
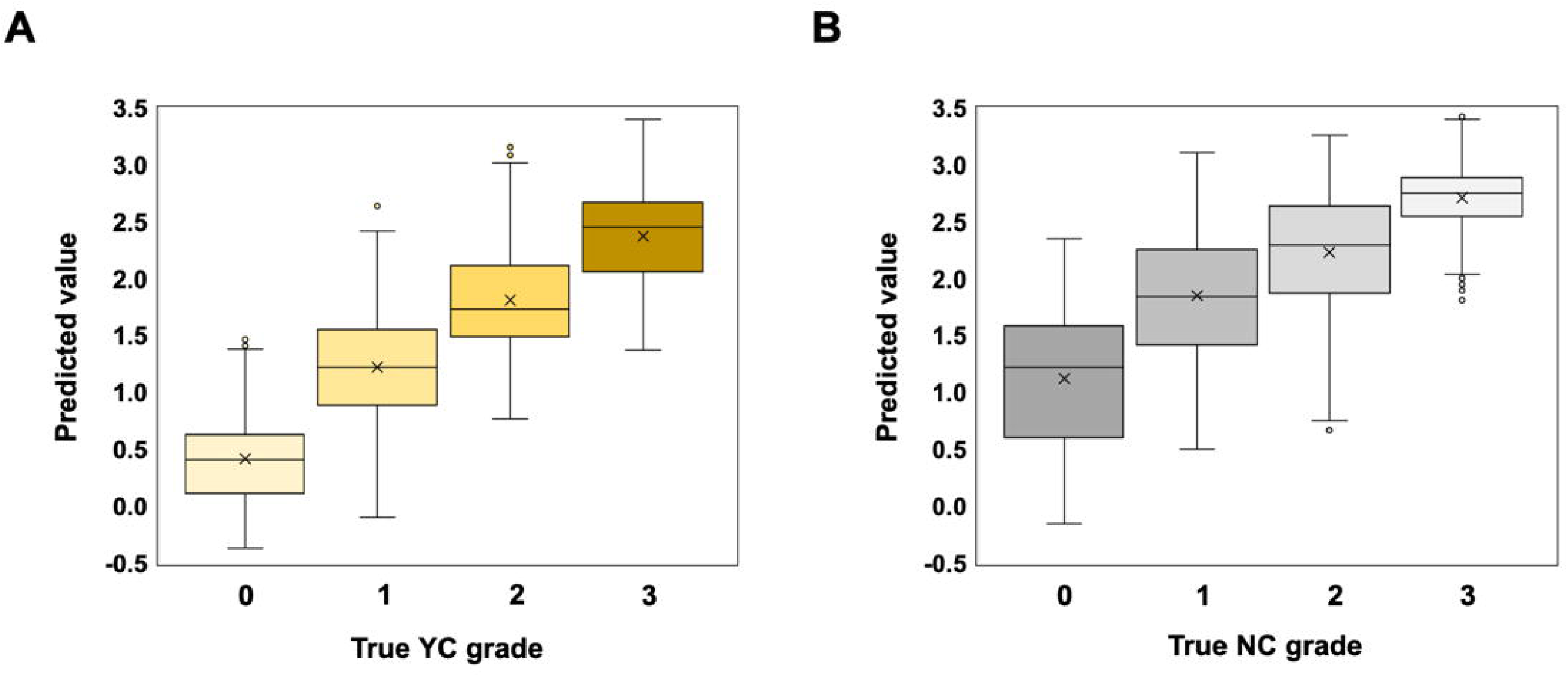
Representative box-whisker plots from the 5-fold cross-validation. The data distribution of the predicted values according to the true values were visualized for yellow grade score (A) and neointimal coverage score (B).

**Fig 5.**
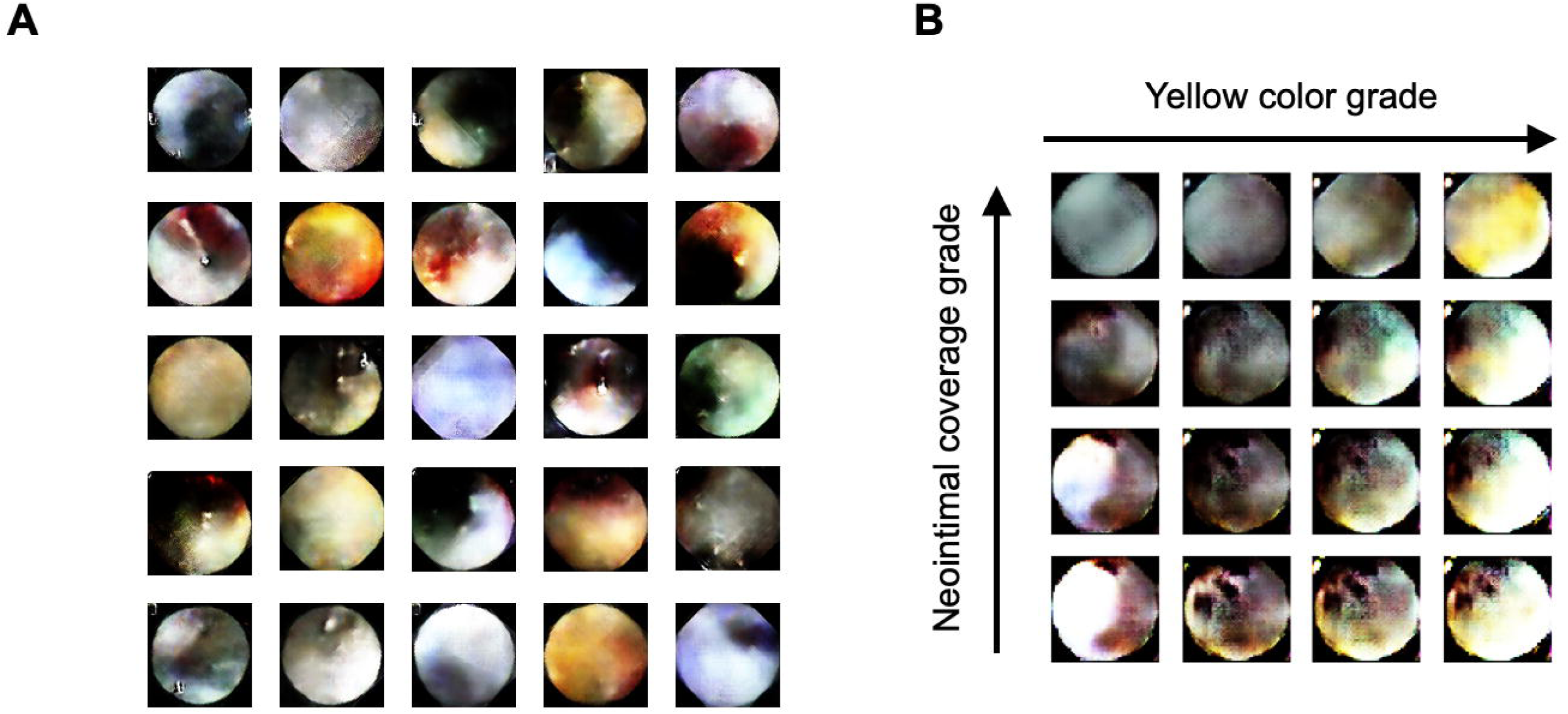
Representative output images. Panel A shows representative output images of the standard GAN model. In panel B, longitudinal axis corresponds to the gradual change of neointima coverage grade and the horizontal axis indicates the grade of yellow color. The thrombus conditions were randomly assigned.

### Incremental effect of GAN-based data augmentation on the prediction model

The baseline performance of the regression model and the effect of GAN-base data augmentation are shown in Table 1. Data augmentation achieved 37.0% increase in the correlation coefficient and reduced 14% in the mean absolute error for the NC grade prediction. In contrast, this method did not significantly affect the performance of YC grade prediction.

**Table 1.** Effect of GAN-based data augmentation on the regression tasks.

|  | YC grade |  | NC grade |  |
| --- | --- | --- | --- | --- |
|  | r-value | MAE | r-value | MAE |
| **Original data**<br>**(n = 864)** | $0.66 \pm 0.03$ | $0.19 \pm 0.01$ | $0.46 \pm 0.02$ | $0.28 \pm 0.01$ |
| **Augmented data**<br>**(n = 1728)** | $0.67 \pm 0.00$ | $0.18 \pm 0.00$ | $0.63 \pm 0.01$ | $0.24 \pm 0.00$ |
| **Fold change** | 1.02 | 0.95 | 1.37 | 0.86 |
| **p-value** | 0.768 | 0.479 | 0.001 | 0.027 |

In the binary classification for the red thrombus, the data augmentation increased 57.9% in F1-score and 5.2% in AUC, owing mainly to the substantial increase of the sensitivity as shown in Table 2.

**Table 2.** Effect of GAN-based data augmentation on the binary classification task.

|  | Precision | Recall | F<sub>1</sub>-score | ROC AUC |
| --- | --- | --- | --- | --- |
| **Original data**<br>**(n = 864)** | $0.81 \pm 0.09$ | $0.26 \pm 0.04$ | $0.38 \pm 0.04$ | $0.77 \pm 0.01$ |
| **Augmented data**<br>**(n = 1728)** | $0.86 \pm 0.04$ | $0.46 \pm 0.04$ | $0.60 \pm 0.05$ | $0.81 \pm 0.00$ |
| **Fold change** | 1.06 | 1.77 | 1.58 | 1.05 |
| **p-value** | 0.614 | 0.008 | 0.006 | 0.002 |

### Performance of the conditional GAN model

Using the conditional GAN architecture, we could control the output images by giving the conditions as shown in Fig 4B. There were significant and strong correlations between given conditions and experts’ score in YC grades (r value = 0.84, p<0.001) and the inter-observer agreement for the grading was r = 0.67 (p<0.001). However, we did not reach agreement on the NC grades in synthesized images (r = −0.31, p=0.011), and there was only a weak correlation between the given conditions and average score (r value = 0.42, p<0.001).

## Discussion

Coronary angioscopy (CAS) is a useful device which allows for the direct visualization of the internal surface of the lumen, providing information about the characteristics of the plaque and thrombus. Previously, we presented evidence that some CAS findings correspond to pathological change (14) and provide an explanation for the other imaging modalities such as optical coherence tomography (2). Angioscopy is also a useful tool to follow up on stent implanted lesions (15). However, due to the limited medical resources, CAS usage is mostly limited for research purposes. Therefore, we believe diagnostic support systems or simulation systems could be beneficial for general cardiologists.

Along with the recent advancement in machine learning algorithms, there are an increasing number of reports concerning the application of artificial neural networks in diagnostic imaging (16). In particular, deep generative modeling has emerged as an effective approach to simulate the complex data structure of medical images (17-20). We recently reported that the deep convolutional encoder-decoder model can be used to reconstruct the apical 2-chamber view in an echocardiogram (21). Considering the DCNN’s ability to expand the potential of diagnostic modalities, we thought that such technologies can be also applicable to broaden the usage of CAS. In this study, we demonstrated the utility of DCNN for the interpretation of CAS views through both prediction tasks and generative modeling.

To begin with, we evaluated the performance of diagnostic prediction models. As previously discussed, one of the principal problems of deep learning in the health science field is the paucity of large training datasets (22). In order to address this issue, we used the MEDLINE database to collect angioscopic images. Because the distribution patterns of the CAS findings were different between the hospital data and literature data, the model’s generalization capacity was expected to increase when the datasets were combined. As a result, the trained models could achieve acceptable performance in YC grade and NC grade prediction. However, the sensitivity in the red thrombus detection model remained relatively low. This might be ascribed to the low incidence of red thrombus in the entire dataset (23) and could be resolved by increasing the sample number or appropriate data augmentation methods.

We next developed a generative model with GAN algorithm to synthesize realistic CAS images. Medical image simulation should be one of the most intuitive applications of generative modeling (24). Recently, Tom F et al. proposed a novel approach to simulate the intravascular ultrasound images with a stacked GAN based framework (25). The authors believe that a simulation model can serve as an aid for doctors to learn rare diseases. Actually, it has been reported that simulation-based training improves cardiology fellows’ skills in cardiac catheterization (26). Since realistic visualization is an essential component of practical simulation (27), our approach to generate CAS images with GAN should be a promising option for the medical simulation method. The medical image synthesis is not limited to educational purposes, but also implicated in the improvement of automatic diagnosis systems. To this date, it has been reported that medical image synthesis by GAN is as effective as a data augmentation method in a classification task (28). Gupta A et al. reported that GAN-based data augmentation could address the class imbalance in the training data and achieved better performance in the bone lesion classification in X-rays (29). Similarly, we could see significant improvement in the performance of the regression model for NC grade and the classification model for the red thrombus by GAN-based image augmentation. Although the validation data size was quite small, our data adds evidence that support the efficacy of this method.

In this study, we also showed the potential of conditional GAN to control the output images according to the CAS findings. With this model, we could change the YC and NC grades of the output image at the same time, by giving the multi-dimensional condition. Although we were not able to control NC grades at this point, this method holds a potential to reduce the burden of annotation tasks for the synthesized images. Aside from that, conditional GAN could be used to further develop a non-invasive virtual coronary angioscopy system (30). As Nishimoto Y et al. reported in 2017, there are some clinical factors which can determine the CAS findings (10). Therefore, if we can select appropriate variables to predict the pathological state from the individual patient data, it might be possible to generate patient-specific virtual CAS images.

## Limitations

This combined dataset may not be reflective of the contemporary cohort of coronary artery disease patients, as the therapeutics for coronary interventions have experienced a drastic change over the last couple of decades. In addition, considerable publication bias is inevitable due to our image collection method. Therefore, it is highly possible that our synthesized images do not reflect the real-world distribution of pathological findings. In fact, we observed higher incidence of red thrombus in the synthesized images than in our hospital data. On the other hand, this publication bias should not have negative influence on the image recognition task, because the collected images were better curated and include more representative findings than our raw data, which can lead to the increased the versatility of our prediction model. The second limitation is the inherent subjective nature of angioscopy itself. Although our 4-point grading scale is widely accepted, there should be a certain degree of intra- and inter-observer variability (31). Quantitative colorimetry is known to be an objective method to assess the plaque color (32), but it was not effective for the YC grade prediction in our data (Supplementary Figure S2). Therefore, our future work should directly focus on the accordance with pathological findings, such as fibrous cap thickness. Finally, the metrics for the generative model in terms of image quality is not complete. Since all the authors were well-versed in angioscopy, Visual Turing test (33) was not applicable this time and we only used Inception score. However, the result of GAN-based data augmentation may indirectly indicate the quality of synthesized images.

## Conclusion

DCNNs are useful in diagnostic prediction, as well as in image generation from the findings in CAS, both of which can contribute to develop a diagnostic support system.

## Data Availability

The publication list where we collected angioscopic images and the source code of the prediction and generative models are available from the public online repository. The de-identified angioscopic images we used in this study are available directly from the corresponding author upon reasonable request.

http://dx.doi.org/10.17632/9dx23j5d64.1

## Acknowledgments

Brandon Shokoples provided constructive comments on the expression in this manuscript.

